# Interplay of Low Serum Irisin Level and Insulin Resistance in Obese Polycystic Ovary Syndrome Patients: Potential Biomarker for Pre-clinical Risk Assessment and therapeutic intervention

**DOI:** 10.1101/2023.11.08.23298152

**Authors:** Kritika Verma, Yogita Rajput, Ruchi Kishore, Pratibha Patel, Ankika Shrivastava, Devpriya Rath, Jagannath Pal, Tripti Nagaria

## Abstract

**Introduction:** Polycystic Ovary Syndrome (PCOS) is the most frequent endocrine syndrome in women of the reproductive age groupassociated with metabolic syndrome and TypeIIDiabetesMalitus (TIIDM). Purpose of the study was to evaluate if level of serum irisin which is involved in energy metabolism could be used as biomarker for identifying high risk category for pre-clinical detection of diabetes in PCOS.

**Methods:** Total 27 obese PCOS, 27 non-obese PCOS, and 27 healthy control (HC: 11 obese and 16 non-obese) study subjects between the age group 15-30 years were recruited in the study. Obese and non-obese category was divided based on BMI. Serum irisin, fasting insulin, fasting glucose and other reproductive hormonal profiles were estimated. Cut offs for serum irisin and insulin level were determined based on levels in healthy control.

**Result:** None of the subject was diagnosed having diabetes at the time of sampling. Both obese and non-obese PCOS showed non-significant lower irisin level than corresponding healthy control (42.27±31.38 vs 51.56±22.7,*P*=0.4 and 34.89±21.58 vs 40.90±16.444,*P*=0.4 respectively). However only obese PCOS showed statistically significant association with lower irisin level when median value of the corresponding healthy control was used as cut off (81.5% vs 45%, *P*=0.047* and 70.3% vs 50%,*P*=0.21 in PCOS vs HC of obese and non-obese group respectively). Segregating PCOS subjects into two category either normal or below the lower limit of normal serum irisin level(mean-1.64*z of corresponding HC), 100% of obese PCOS subjects having irisin below the lower normal limit showed insulin resistance (>mean+2SD of non obese HC) while the same in obese PCOS subjects having irisin level above the lower limit was only 20% (*P*=0.006). However no such association was observed in non-obese PCOS subjects when similarly categorized based on irisin level (33% vs 22% respectively, *P*= 1).

**Conclusion:** The result suggests that obese PCOS with serum irisin level below lower limit of normal range could be segregated as high risk category for closely follow up to detect incidence of diabetes in early time point or taking any preventive measure in early stage. A prospective study with larger sample size is required to substantiate the observation of the pilot study.

## 1. Introduction

Polycystic ovary syndrome (PCOS) is a one of the common endocrine syndrome in women of reproductive age groupwith an occurrence of around 4 to 10% all over the world^1^.Prevalence of PCOS in India is high and it was close to 10% using Rotterdam’s criteria and AES criteria whereas 5.8% by NIH criteria^2^. PCOS manifests as menstrual dysfunction, infertility, hirsutism, acne, and obesity. It defines a condition where at least one ovary has an ovarian volume greater than 10 mL and at least one ovary has an estimated ten small cysts, with diameters ranging from 2 to 9 mm, develop^3^. It is usually only diagnosed when complications develop that significantly reduce a patient’s quality of life like hair loss, alopecia, acne, and infertility-related problems ^4,5^.

PCOS starts with menstrual irregularity in teenager to infertility in young adult and to end with metabolic syndrome in elderly women. Women with PCOS are often insulin resistant, increasing their risk for type II Diabetes Mellitus (T2DM). It is an Insulin resistance (IR) state like obesity and T2DM. Women with PCOS have three-fold increased risk for impaired glucose tolerance (IGT) and increased risk for T2DMin contrast to women from the general population. As per previous observation, more than half of women with PCOS develop T2DMby age 40. Forslund et al 2020^6^ also reported that Obese PCOS women in the mid-fertile years are prone to develop T2DM. Early diagnosis through closely follow-up of high risk PCOS patients could prevent late stage complications from T2DM and other metabolic syndrome.

In normal physiology there are several mechanisms to keep the peripheral tissue sensitive to insulin. One of the newly discovered peptide hormone Irisin discovered by Bostrom et al.^7^ whichhelps actually reducing the insulin resistant stage, increasing the insulin production from pancreas as well as reduce the peripheral insulin resistance by white adipose tissue (WAT) to brown adipose tissue (BAT). Irisin is a myokine, produced as proteolytic cleaved product of Fibronectin domain containing protein 5 (FNDC5) in the outer membrane of muscle cell. In normal condition it is present in dormant and inactive stage. Exercise and other unknown factors cause this protein to be split from the N-terminal site and secrete soluble protein called Irisin, its peptide length 112 amino acid (Bostrom et al. 2012^7^& Schumacher et al. 2013^8^). It also facilitates the insulin production by the act on islet cells of pancreas therefore it improves glucose tolerance. It is also responsible for reducing obesity by calorie utilization. So impaired irisin level could be considered to be the future target for diagnosis as well as therapies for metabolic disorder.

As per previous reports, it has been demonstrated that irisin concentration is elevated in PCOS but shows no correlation between body mass index (BMI) with irisin level (Sharan et al 2021^9^). However, few studies have found that adult women with PCOS had either significantly lower mean serum irisin concentrations, or although decreased (Abali 2016^10^) as well as they did not find any correlation with hormonal profile of PCOS patients, there was no difference when compared to controls (Gao et al 2016^11^). Thus,There is conflicting evidence on serum irisin level in women with PCOS from study to study. While some study reported elevated irisin in PCOS^12-16^, others observed either no changes or decrease level of irisin^10,11^.In obese patients due to high BMI there is relative insulin resistance and to compensate that there is increase irisin demand. If insulin and irisin balance is deranged it may precipitate TIIDM. However none of the previous study did not established the direct interplay of insulin and irisin level in obese and non obese PCOS patientsto identify the specific high risk group. In this study we have evaluatedthe interplay of irisin and insulin level in obese and non-obesePCOS patientsusing a cut off irisin and insulin level in normal healthy control.

## 2. Research Design / Methodology

This case–controland cross sectional study was conducted in the Department of Obstetrics and Gynecology and Multi disciplinary Research Unit (MRU)of TertiaryCareHospital Dr.B.R.A.M. Hospital under Pt. J.N.M.Medical College Raipur, Chhattisgarh, India, following the approval by institutional ethical committee. This studyinvolves two group i.e. PCOS and healthy controls. Inclusion criteria for PCOS patients to be the presence of any 2 of the following 3 criteria on the basis of revisedRotterdam European Society of Human Reproduction and Embryology (ESHRE)/American Society of Reproductive Medicine (ASRM)^5^

Exclusion criteria: More than 45 years were the exclusion criteria for both groups, patients suffering from Cushing’s syndrome, thyroid dysfunctions, androgen-secreting tumor, enzyme deficiency (21-hydroxylase in particular), decreased ovarian reserve (primary ovarian insufficiency), or type 1 or type 2 diabetes were excluded. None of the subjects will be taking any medication for at least 3 months before the study. Smoking and alcohol consumption should be the exclusion criteria for both group case as well as control.

Healthy controls: 1. Age and sex matched with PCOS patients, 2. non-obese 3. Not known medical disorder 4. Non hirsutism 5.Similar socioeconomic status.

PCOS patients were divided into two subgroup, first PCOS with obesity (PCOS+obese) and second group is lean PCOS patients (PCOS+non-obese).

### Sample collection

Participants of both study and control groups were tested within 3^rd^ to 5^th^ days of menstruation and the fasting blood sample collection timing will be in the morning hours approximately 9 to 10 am.

### Anthropometric measurements

Body Mass Index (BMI) were assessed by weight, hight and waist circumference for all.

### Estimation of Hormonal and biochemical markers

Plasma sample werecollected for different biochemical analysis *i*.*e*. fasting glucose (fGlu), fasting insulin (fI), HbA1c, Free fatty acid (FFA), total cholesterol (TC), high-density lipoprotein cholesterol (HDL-C), low-density lipo-protein cholesterol (LDL-C), and triglyceride (TG). Among hormones luteinizing hormone (LH), follicle-stimulating hormone (FSH), testosterone (T) and progesterone (Prog), thyroid stimulating hormone (TSH), pro-lactine (PRL), free testosterone (fT), total testosterone levels, Dehydroepiandrostenedione sulfate (DHEA-S) and the sex hormone binding globulin (SHBG) will be estimated.

### Estimation of Irisin

commercially available ELISA kit was used for the estimation of Irisin level in serum sample of PCOS patients as well as healthy groups.

### Statistical analysis

Mean, standard deviation, and Pearson correlation were performed. Comparison of mean, significance ofPearson’s correlation, significance of associationFisher exact probability due to small sample size analyzed using the following webbased software: MEDCALC https://www.medcalc.org/calc/comparison_of_proportions.php^17^.P-values ≤ 0.05 were considered statistically significant.

## 3. Results

### 3.1 Demographic and clinical profile of obese and non obese PCOS patients compared with their corresponding healthy controls

Total54female cases of PCOS and 27 healthy subjects aged between 18 to 45 years were participated in this study. Out of54 PCOS cases, 27 were Obese (BMI >24.9 kg/m^2^) and 27 were non obese(BMI <24.9 kg/m^2^). Among 27 healthy control 11 were obese and 16 were non-obese.Mean age of cases and control were 24.67± 5.03 year and 26.74±3.64 yearrespectively and the difference was not statistically significant. In our study most of the subjects were belonged to the age group 21-30 years (66.6%). Detail age and demographical distribution is given in table 1. Most of the PCOS cases reside to urban area (79.6%)however maximum belongs to upper Middle Class (48.1%) wherein the occurrence are prevalence in students (44.4%).

**Table 1:**
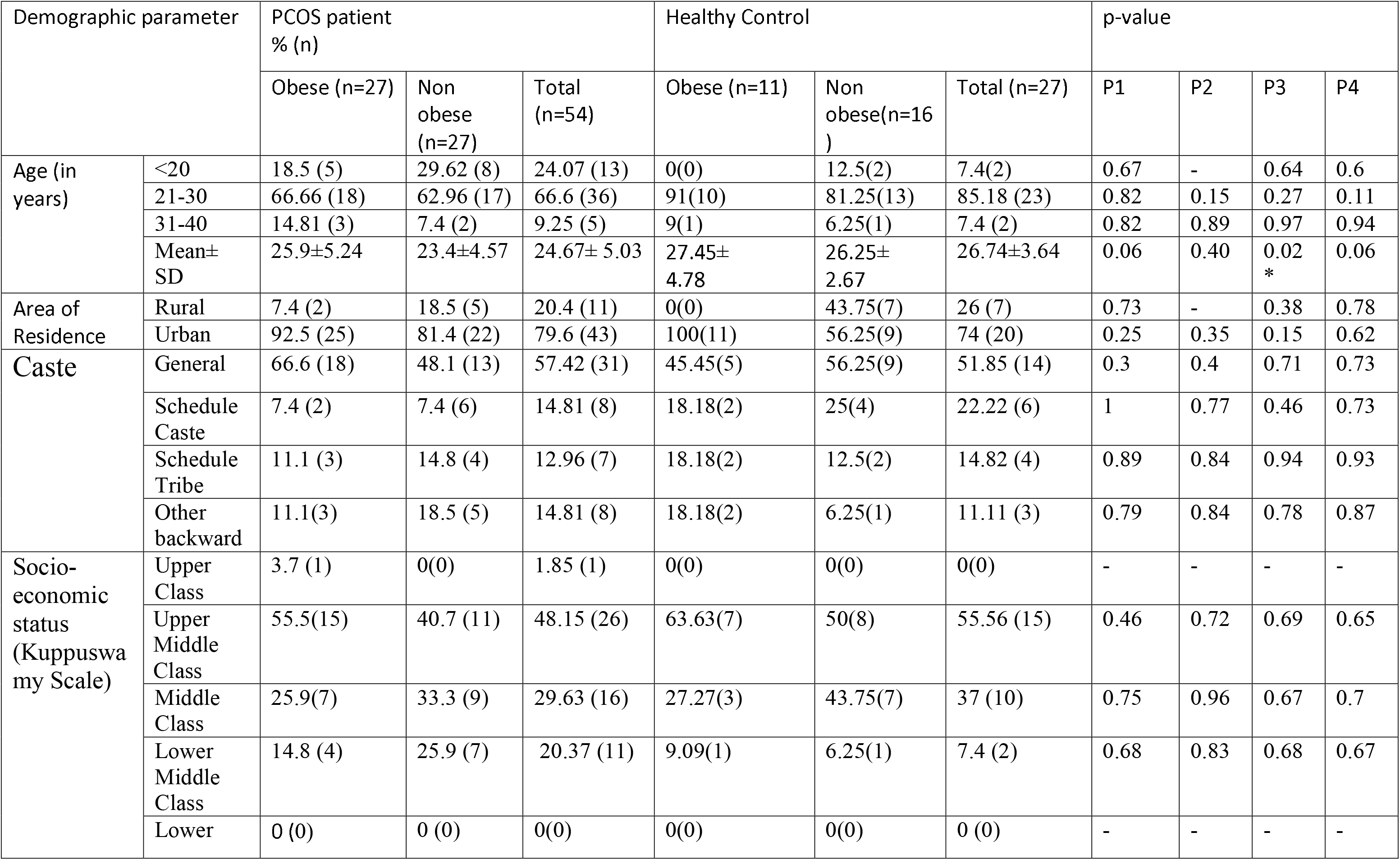
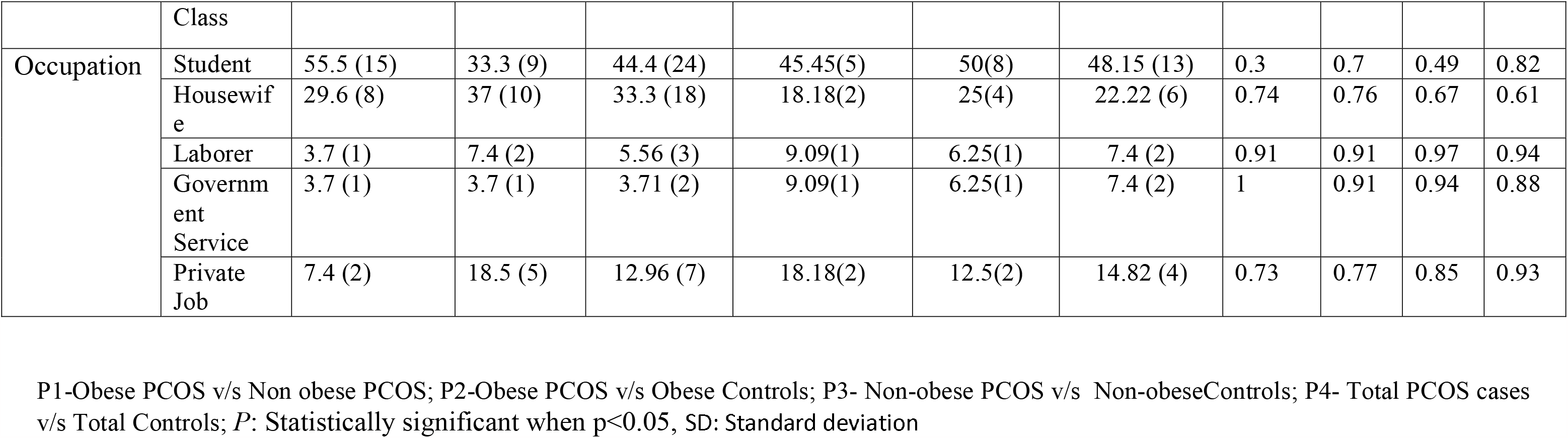
Socio-Demographic and Anthropometric Profile of obese and non obese PCOS patients compared with healthy controls.

Maximum cases had weight in the range of 50-59.9 kg (42.5%).Mean weight was found more in obese and difference was statistically significant (P: 0.04). No significant difference was found on comparing height amongst PCOS cases (P: 0.9).Difference of weight between obese and nonobese PCOS was significant P<0.0001.Detail profile of Waist Hip Ratio(WHR), Abdominal Circumference (AC) and BMIin Obese and non-obese PCOS cases and control are given in table 2.Most common menstrual abnormality, noted in cases was amenorrhea (38.8%) followed by oligomenorrhea (25.9%) (table 2).Maximum cases (35.1%) had menarche by the age of 13 years.Acne was found in 42.5% cases with PCOS. 22.2% obese PCOS cases presented with acanthosis nigricans.

**Table 2:**
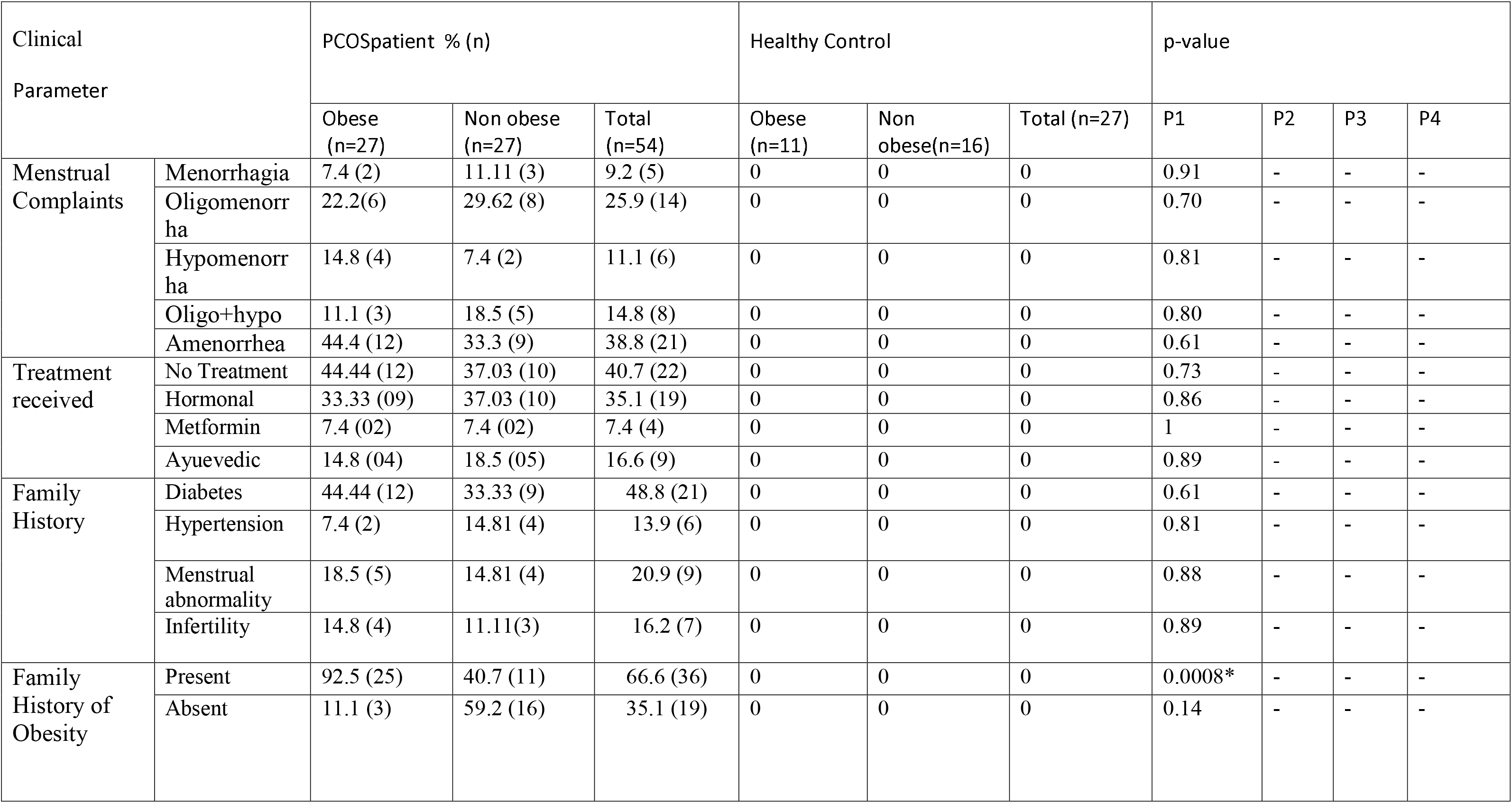
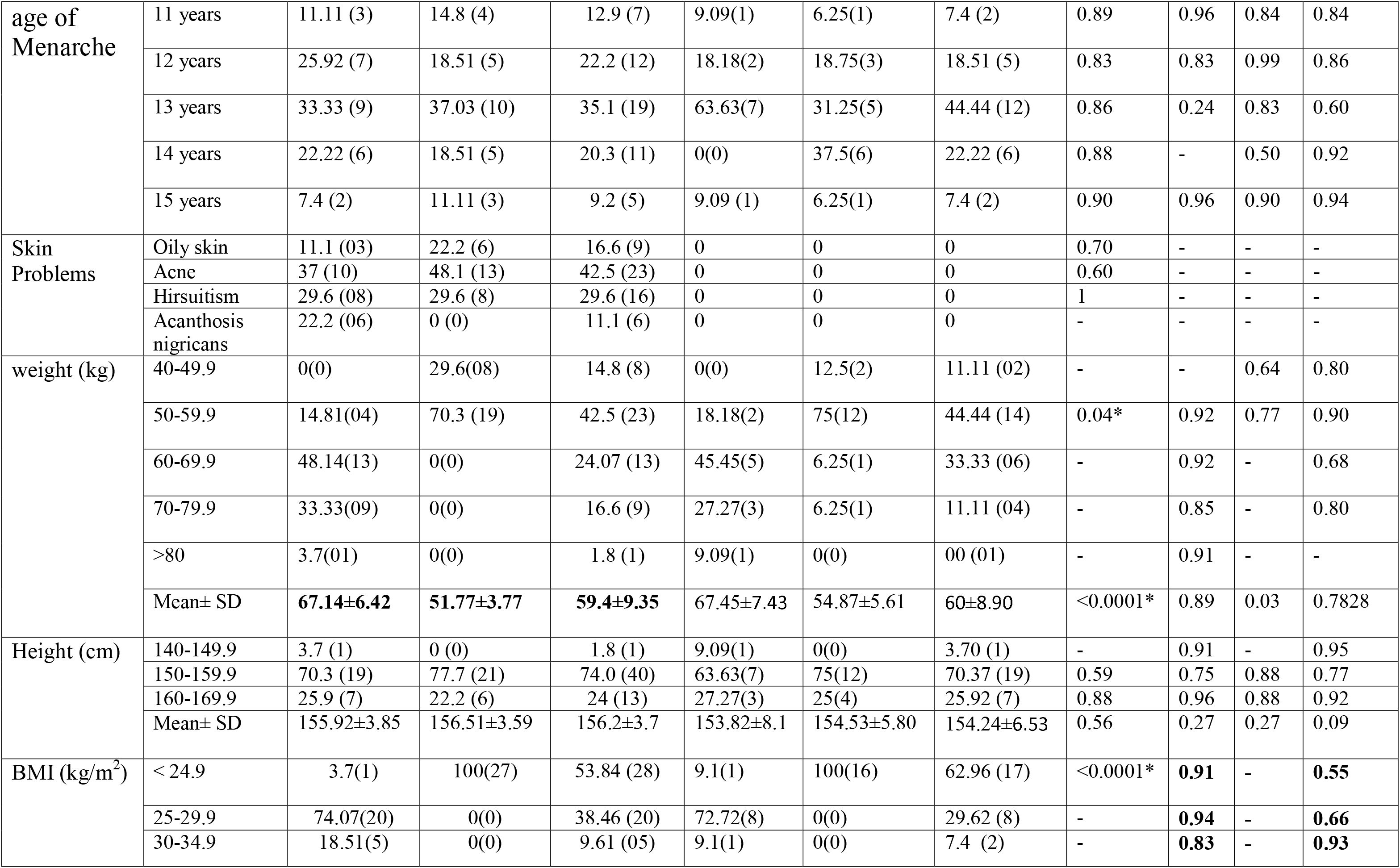
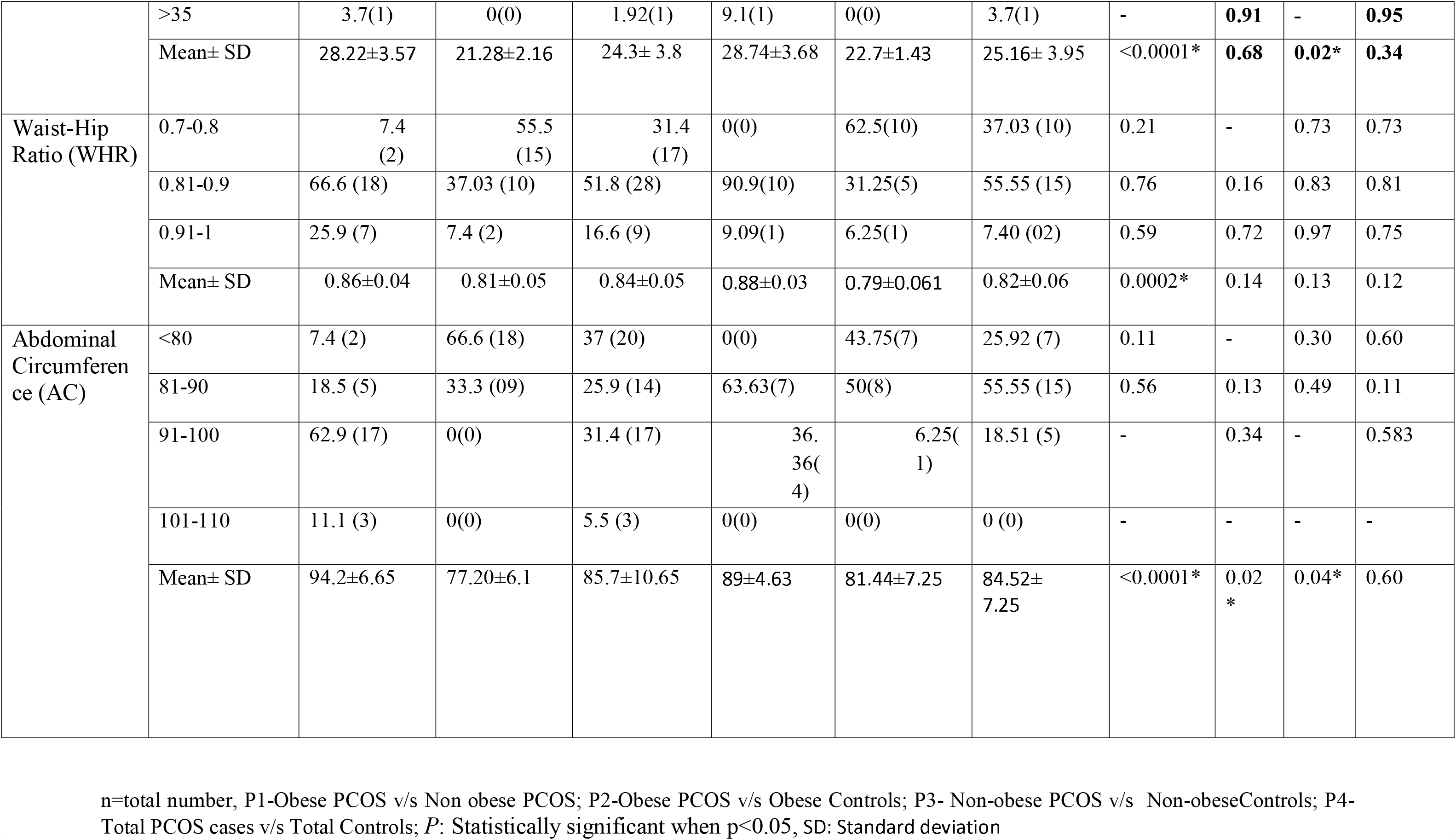
Clinical profile of obese and non obese PCOS patients compared with healthy controls.

### 3.2 Reproductive hormonal profile of obese and non obese PCOS patients compared with their corresponding healthy controls

Serum level of Ovarian hormonal profile of PCOS patientgroups (Testosterone, estrogen, progesterone)were lowered whereas level of gonadotropic hormones (Luteinizing hormone (LH), follicle-stimulating hormone (FSH))wereelevated compared to their corresponding healthy controls (Table 3).LH:FSH ratio was significantly increased in PCOS patients (1.8) relative to healthy control (0.97), (P: <0.0001) as expected.however, in obese cases the ratio washighercompared to non obese (P: 0.0001). Significant difference was found between cases and controls on comparing serum prolactin (P:0.001)whereas non significant difference was observed in Thyroid-stimulating hormone (TSH) levels.

**Table 3:**
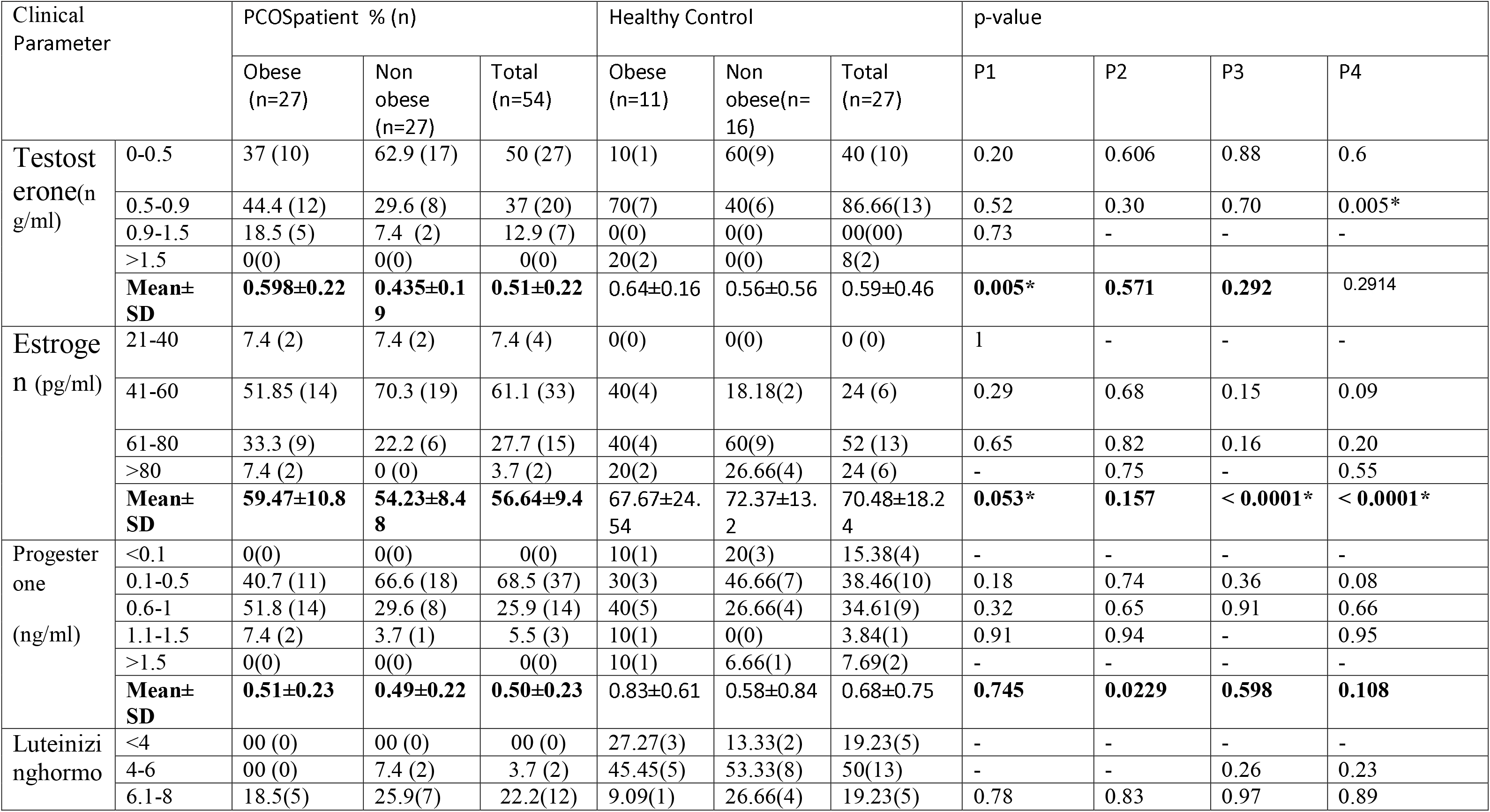
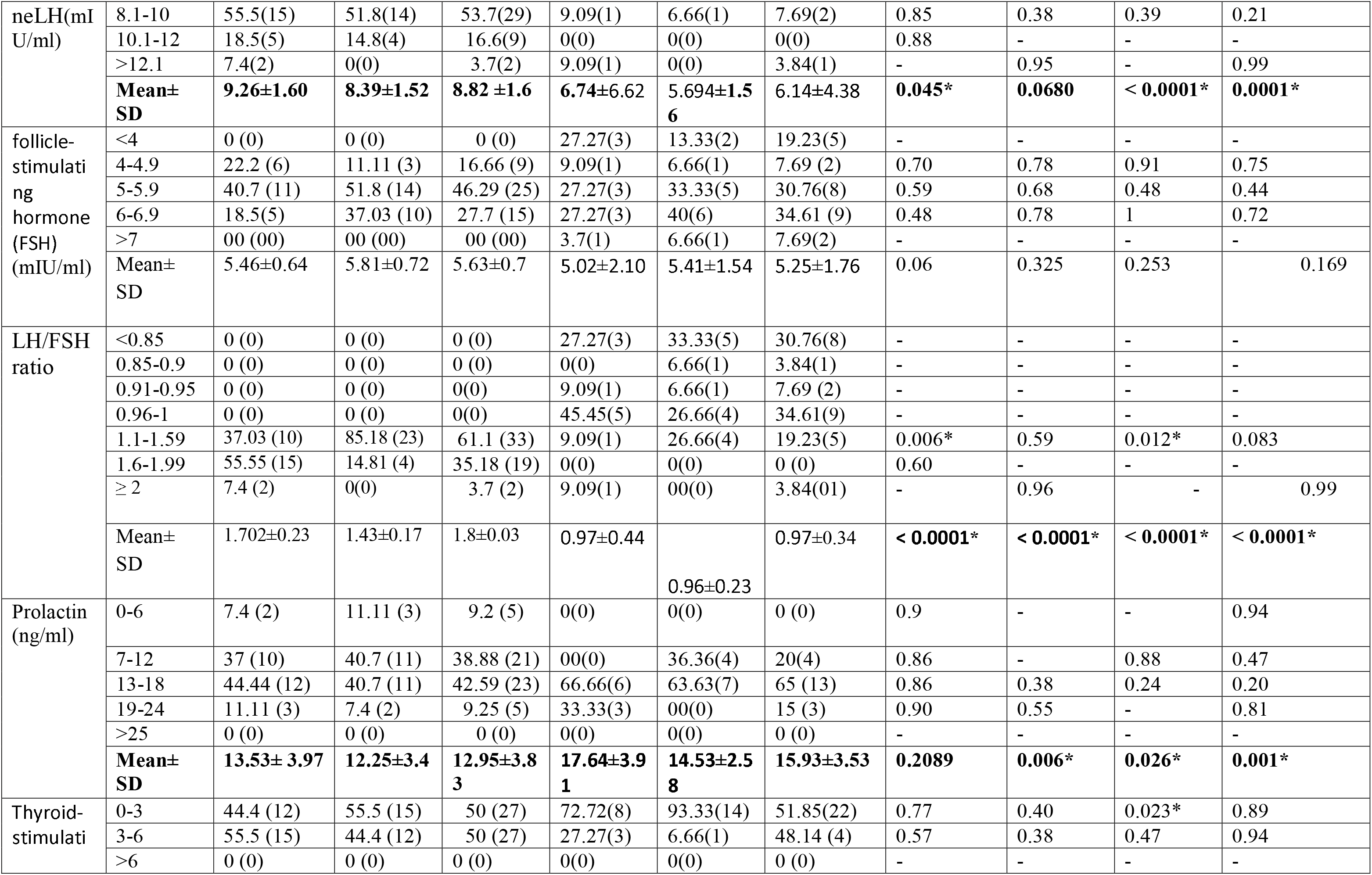
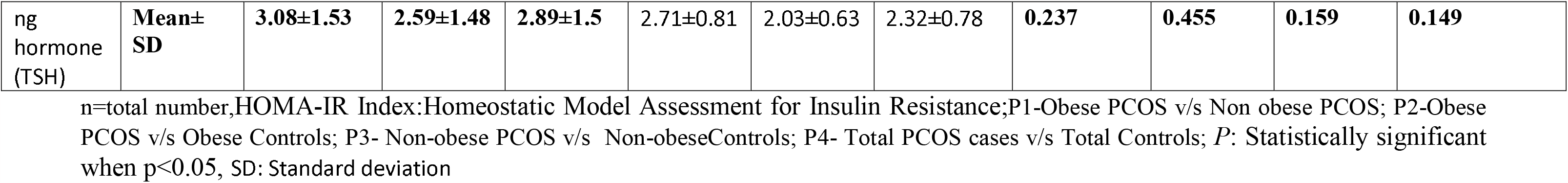
Hormonal profile ofobese and non obese PCOS patients compared with healthy controls.

### 3.3 low serum irisin level is associated with obese PCOS patients compared to corresponding controls

Circulating irisin is one of key player in glucose metabolism andinsulin sensitivity.As PCOS patients are prone to develop type 2 diabetes,we evaluated if there is any alteration of irisin levels in PCOS patients compared to corresponding healthy controls. Overall serum irisin levels were detected lower in PCOS cases (38.58±26.94 ng/ml) compared to the healthy control group (45.24±19.56 ng/ml) (table 4), however the difference was not statistically significant (P=0.347). when the subjects were categorized into obese and non-obese groups, expectedly mean irisin levels in obese PCOS and obese healthy control were higher than in corresponding non-obese groups without any statistical significance (42.27±31.38 and 51.56±22.7 Vs 34.89±21.58 and 40.90±16.44 respectively). Relatively more elevated levels of irisin in obese subjects indicate higher need for balancing energy metabolism. This indicate a imbalance of irisin level to meet the need as per the body mass index in PCOS compared to healthy control.When we make the median value of irisin levelas cut off in obese/non-obese healthy control,out of 27 obese patients, 81.5% showed irisin level below the corresponding cut-off(<50ng/ml), as shown in Table 5.The associationof low irisin in obese PCOS compare to corresponding obese healthy control werestatistically significant (odd ratio 95% CI: 1.1-24.5;P = 0.033* and Fisher’s exact probability P = 0.047*). However among the non-obese subjects, as per our criteria (median value cut off: 39.7∼40 ng/ml), 70.3% (n = 19) of PCOS patients had lower than the cut-off but the association was notstatistically significant.

**Table 4:** Serum irisin concentration in PCOS cases and Control (Parametric analysis)

| Irisin concentration (ng/ml) | PCOS |  |  | Controls |  |  |
| --- | --- | --- | --- | --- | --- | --- |
|  | Total<br>(n=54) | Obese<br>(n=27) | Non obese<br>(n=27) | Total<br>(n=27) | Obese<br>(n=11) | Non obese<br>(n=16) |
| Median | 32.75 | 35.29 | 32 | 42.04 | 50.3 | 39.7 |
| Mean±SD | 38.58±26.94 | 42.27±31.38 | 34.89±21.58 | 45.24±19.56 | 51.56±22.7 | 40.90±16.44 |
| P-value<<br>(0.05) | 0.347<br>(Vs HC) | 0.41<br>(Vs HC Obese) | 0.40<br>(Vs HC Non Obese) | 0.30 (HC Obese vs non obese) |  |  |
|  |  | 0.318 (PCOD Obese vs non obese) |  |  |  |  |

P: Statistically significant when $p < 0.05$ ; HC=Healthy Control, SD: Standard deviation

**Table 5:** Association of low serum irisin level in PCOS patients (Non parametric analysis) in obese and non-obese patients compared to healthy control.

| Risk Factor:<br>Serum<br>Irisin(ng/ml) in<br>Obese & non-obese | | PCOD<br>% of total (n) | HC<br>% of total (n) | Odd ratio<br>OD (95%CI)<br>$P^{***}$ | Fisher exact<br>probability:<br>$P^{***}$ |
| --- | --- | --- | --- | --- | --- |
| Obese | <50* | 81.5 (22) | 45.5 (5) | 5.3(1.1-24.5) | $P= 0.047^*$ |
| | >50 | 18.5 (5) | 54.5 (6) | $P= 0.033^*$ | |
|  | Total | 27 | 11 |  |  |
| Non-Obese | <40** | 70.3 (19) | 50 (8) | 2.37 (0.66-8.56) | $P=0.209$ |
| | >40 | 29.6 (8) | 50 (8) | $P=0.186$ | |
|  | Total | 27 | 16 |  |  |

\*Median value of irisin in Obese Healthy control(HC) = 50.3ng/ml; \*\* Median value of irisin in Non-Obese Healthy control = 39,7ng/ml; P: Statistically significant when $p < 0.05$ .

### 3.4 Correlation between Serum irisin level with clinical manifestation, Reproductive hormonal profile as well as demographic profile

The correlation of serum irisin level with serum reproductive hormonal profile as well as demographic profile are depicted in Table S1 and figure S1. Irisin level was Significantly correlated with onlyserum testosterone in obese PCOS patients (R^2= 0.4194, P= 0.002*) and with serum progesterone in non obese PCOS subjects (R^2= 0.9436; P=<0.0001*).

### 3.5 Profile of glucose homeostasisin PCOS patients compared with their correspondingHealthy controls

Hyperglycemia, hyperinsulinemia, insulin resistance, etc. are indications of metabolic disturbances that are commonly observed as metabolic syndrome in PCOS. In our study we did not find any difference in fasting glucose levelin PCOS patients compared to corresponding healthy controls (92.99±9.74 vs 96.63±6.61 and 87.44±7.46 vs92.5±10.08 in obese and non-obese PCOS vs corresponding healthy control respectively). There was no overall significant mean difference of fasting insulin levelswas observed in Obese PCOS compared to the corresponding healthy control (18.12±6.34 vs17.11±5.44, P=). Similarly no overall significant mean difference of fasting insulin levels was observed innon-obese PCOS compared to the corresponding healthy control (15.15± 3.55 vs9.78±4.97 P=).However when we correlated fasting glucose and insulin as a HOMA-IR indexin our study group, obese (4.30 ±1.96) and nonobese (3.32±1.01) PCOD patients as well as obese healthy control (4.17±1.50) their HOMA-IR index were > than 2.9 as compared to corresponding nonobese healthy control (2.19±1.12) which is the indication of significant insulin resistance state (table 6). When we observed the frequency distribution of fasting insulin response in obese and non-obese PCOS patients compared with corresponding healthy controls shows peck of obese PCOS patients were inclined toward extreme right side suggesting more association with insulin resistant (figure 1).

**Table 6:** Profile of glucose homeostasis in PCOS patients compared with their corresponding Healthy controls.

| Clinical Parameter |  | PCOD patient % (n) |  |  | Healthy Control |  |  | p-value |  |  |  |
| --- | --- | --- | --- | --- | --- | --- | --- | --- | --- | --- | --- |
|  |  | Obese (n=27) | Non obese (n=27) | Total (n=54) | Obese (n=11) | Non obese(n=16) | Total (n=27) | P1 | P2 | P3 | P4 |
| Fasting Insulin levels (µU/ml) | 0-9.9 | 7.4 (2) | 11.11 (3) | 9.25 (5) | 11.11(1) | 50(7) | 34.78 (8) | 0.90 | 0.92 | 0.27 | 0.32 |
|  | 10-19.9 | 51.85 (14) | 85.18 (23) | 68.51 (37) | 55.55(5) | 50(7) | 52.17 (12) | **0.03\*** | 0.89 | **0.05\*** | 0.41 |
|  | 20-29.9 | 37.03 (10) | 3.7 (1) | 20.3 (11) | 33.33(3) | (0) | 13.04 (3) | 0.52 | 0.91 | - | 0.78 |
|  | 30-39.9 | 7.4 (2) | 00 (00) | 3.7(2) | 0(0) | (0) | 00 (0) | - | - | - | - |
|  | **Mean±SD** | **18.12±6.34** | **15.15± 3.55** | **16.6±5.3** | **17.11±5.44** | **9.78±4.97** | **12.65±6.22** | **0.03\*** | 0.64 | **0.0002\*** | **0.004\*** |
| Fasting Blood glucose (FBG)(mg/dl) | <80 | 18.51 (5) | 18.51 (5) | 18.5 (10) | 9.09(1) | 18.75(3) | 14.81 (4) | 1 | 0.83 | 0.99 | 0.87 |
|  | 81-90 | 29.62(8) | 51.85 (14) | 40.7 (22) | 27.27(3) | 25(4) | 25.92 (7) | 0.32 | 0.94 | 0.35 | 0.48 |
|  | 91-100 | 29.62(8) | 18.51 (5) | 24 (13) | 18.18(2) | 43.75(7) | 33.33 (9) | 0.67 | 0.75 | 0.38 | 0.64 |
|  | >100 | 22.22(6) | 11.11(3) | 16.6(9) | 45.45(5) | 12.5(2) | 25.92 (7) | 0.70 | 0.43 | 0.96 | 0.65 |
|  | **Mean±SD** | **92.99±9.74** | **87.44±7.46** | **89.7±9.48** | **96.63±6.61** | **92.5±10.08** | **94.18±10.88** | **0.023\*** | **0.26** | **0.06** | **0.06** |
| HOMA-IR Index | 0-1.8 | 0(0) | 11.11 (3) | 7.4 (4) | 11.11(1) | 42.85(6) | 30.43(7) | - | - | 0.36 | 0.40 |
|  | 1.9-2.8 | 33.3 (9) | 25.9 (7) | 27.7 (15) | 00(0) | 28.57(4) | 17.39(4) | 0.76 | - | 0.93 | 0.68 |
|  | 2.9-3.8 | 18.5 (5) | 25.9 (7) | 22.2 (12) | 18.18(2) | 14.28(2) | 17.39(4) | 0.77 | 0.99 | 0.74 | 0.84 |
|  | 3.9-4.8 | 11.11(3) | 29.6 (8) | 20.3 (11) | 33.33(3) | 14.28(2) | 21.73(5) | 0.54 | 0.55 | 0.67 | 0.94 |
|  | 4.9-5.8 | 14.8 (4) | 7.4 (2) | 11.1(6) | 33.33(3) | 00(0) | 13.04(3) | 0.81 | 0.59 | - | 0.93 |
|  | 5.9-6.8 | 11.1 (3) | 0(0) | 5.5(3) | 00(0) | 00(0) | 00(0) | - | - | - | - |
|  | 6.9-7.8 | 3.7 (1) | 0 (0) | 1.8 (1) | 00(0) | 00(0) | 00(0) | - | - | - | - |
|  | >8 | 7.4 (2) | 0 (0) | 3.7 (2) | 00(0) | 00(0) | 00(0) | - | - | - | - |
|  | **Mean±**<br>**SD** | **4.30 ±1.96** | **3.32±1.01** | **3.8± 1.63** | **4.17±1.5**<br>**0** | **2.19±1.1**<br>**2** | **2.96±1.60** | **0.02\*** | **0.84** | **0.001\*** | **0.03\*** |

n=total number,HOMA-IR Index:Homeostatic Model Assessment for Insulin Resistance;P1-Obese PCOS v/s Non obese PCOS; P2-Obese PCOS v/s Obese Controls; P3- Non-obese PCOS v/s Non-obeseControls; P4- Total PCOS cases v/s Total Controls; *P*: Statistically significant when $p < 0.05$ ,SD:Standard deviation

**Figure 1:**
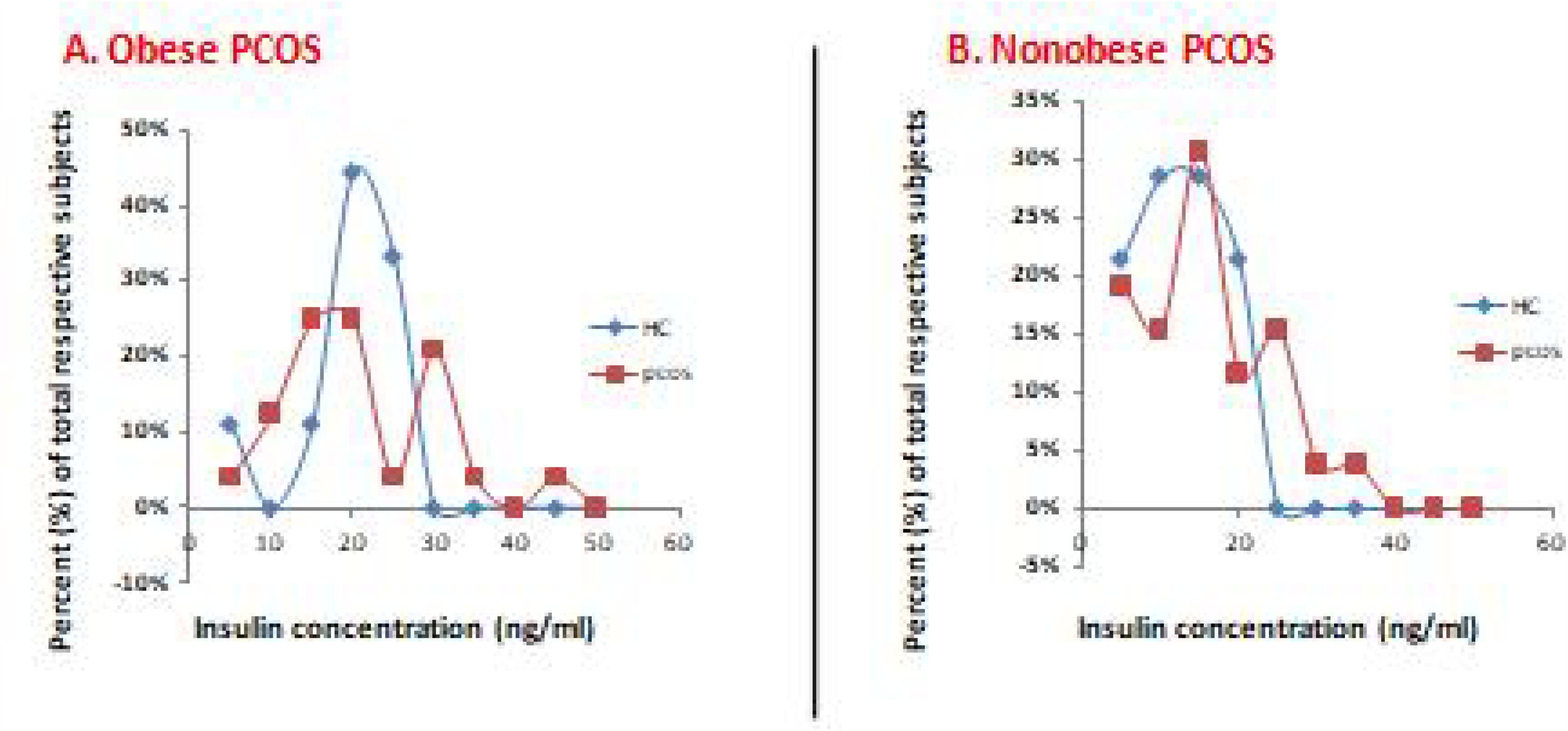

**Figure 3.** Frequency distribution of fasting insulin in obese and non obese PCOS patients compared with corresponding healthy control. (A)Frequency distribution of fasting insulin level of Obese PCOS patients and obese healthy control. (B) Frequency distribution of fasting Insulin level of non obese PCOS patients and non obese healthy control. HC: healthy control;

### 3.6 Increasedlevel of fasting insulinis associatedwith low serum irisin level in Obese PCOS patients

As insulin sensitivity is regulated by irisin,association of insulin response to low irisin level were evaluated in PCOS patients compared to corresponding healthy controlsparametrically (Table 7) as well as non-parametrically (table 8).For defining significantly low irisin level we separately determined the one sidedZ score for 95% percentile (1.645 * SD) of obese and non-obese healthy control (Mean-1.64*SD :14.3 & 13.9ng/ml respectively).One sided Z score was taken because only low irisin level have the pathological interpretation. As both showed very close value, the cut off 14 ng/ml was taken as lower limit of normal range for all PCOS cases. Any value below that limit was considered significantly low irisin level.

**Table 7:** Association of insulin response with serum irisin level in PCOS patients (parametric analysis)

| Irisin concentration (ng/ml) </>14* | PCOD in obese |  | PCOD in nonobese |  |
| --- | --- | --- | --- | --- |
|  | <14 | >14 | <14 | >14 |
| Insulin level (Mean±SD) | 27.525±2.12 | 16.107±32.27 | 15.213±2.74 | 13.00±21.18 |
| P< (0.05) | 0.000058*** |  | 0.0000079*** |  |

\*Cut off value of irisin:14ng/ml [Lower limit of irisin for normal range:one sided Z score for 95% percentile 1.645 \* SD of healthy control (Mean±SD\*1.64)= 14.3 & 13.9 for obese and non-obese respectively, cut off 14 is taken for all PCOD cases], n: Number of subjects, CI: Confidence Interval, \* Pstatistically significance at P<0.05, SD: Standard deviation

**Table 8:** Association of insulin response with serum irisin level in PCOS patients (Non parametric analysis)

| Risk Factor in PCOD: Serum Irisin </>14* |  | Insulin Resistance >19.7 [% (n)] | Insulin Sensitive <19.7 [% (n)] | Total | Odd ratio OD (95%CI) P** | Fisher exact probability: P** |
| --- | --- | --- | --- | --- | --- | --- |
| Obese PCOD | <14 | 100 (4) | 0 (0) | 4 | 33(1.48 to 734.11), P= 0.027 | P=0.00659* |
|  | >14 | 20 (4) | 80 (16) | 20 |  |  |
| non-obese PCOD | <14 | 33(1) | 67(2) | 3 | 1.8(0.13 to 24.16) P= 0.65 | P=1.0 |
|  | >14 | 22(5) | 78 (18) | 23 |  |  |

\*Cut off value of irisin:14ng/ml [Lower limit of irisin for normal range:one sided Z score for 95% percentile 1.645 \* SD of healthy control (Mean±SD\*1.64)= 14.3 & 13.9 for obese and non-obese respectively, cut off 14 is taken for all PCOD cases]; Cut off value for insulin resistance: 19.7(mIU/ml)[ upper limit of normal range in non obese subjects (Mean+2SD)]; P: Statistically significant when p<0.05.

In obese PCOS patient group, the subjects having irisin level below 14 ng/ml, the mean fasting insulin level was significantly higher than those subjects having irisin level in normal range (27.525±2.12 vs 16.107±32.27 respectively, P=0.000058). In non obese PCOS subjects also, significant higher mean fasting insulin was observed in low irisin sub-group compared to sub group having normal range of irisin level (15.213±2.74vs13.00±21.18, P=0.0000079) shown in table 7.

For non parametric association of low level of irisin with insulin resistance, ‘insulin resistance’in our study was defined as the higher fasting insulin level than upper cut off limit of normal range innon-obese healthy control group (mean +2SD:19.7μU/ml).Our result showed 100%(4/4) of obese PCOS patients having low irisin level (<14ng/ml) have insulin resistance as defined above (>19μU/ml fasting insulin) where as only 20 % (4/20)obesePCOS patients having irisin level in normal range (>14ng/ml) have fasting insulin >19μU/ml. The association of insulin resistance with low irisin level showed statistically significant (odd ratio: 33, 95%CI: 1.48 to 734.11), P= 0.027; Fisher exact test: P=0.00659). In contrastonly 33% of non-obese PCOS patients having low irisin level (<14ng/ml) were found to have insulin resistance(>19mμIU)compared to 22 % (4/20) ofnon-obese PCOS patients having irisin level in normal range (>14ng/ml) were showedinsulin resistance without any statistical association(odd ratio: 1.8, 95%CI: 0.13 to 24.16), P=0.65; Fisher exact test: P=1) (table 8).

## Discussion

The present study is to find the levels of serum irisin in Polycystic ovarian syndrome cases and to find its association with insulin resistance. There is very inconsistent observation of fasting Serum irisin level in PCOS. While some earlier study showed increase irisin level^12-16^in PCOS where as other studies showed either no increase or decrease level of irisin in PCOS patients^10,11^. In most of those studies obese and nonobese control and PCOS groups were not reported separately^12-16^. Li et al.2015^13^, had reported overall increase fasting irisin level than the normal woman control group. However in the same study increase irisin was observed in both obese PCOS and obese normal compared to corresponding non obese groups. Irisin level is increased in obesity to overcome the relative insulin resistance. As irisin level is increased with BMI, it should be evaluated in obese (PCOS & normal) and non obese (PCOS & normal) groups separately to understand the dysregulation of irisin in PCOS, eliminating confounding effect of BMI.So to address the discrepancy we evaluated irisin level of PCOS with the obese and non obese compared to corresponding healthy control groups and correlate with other reproductive hormonal level.

In our study we observed expectedly ovarian hormonal level (testosteronestrogen, progestron)were decreased whileLH/FSH)ratio were increasedin PCOS compared to corresponding healthy control as described before^9,18^.

Though there is low mean irisin level in obese and non obese PCOS than corresponding healthy control however the difference was not statistical significant. This might be due to low sample size &/or wide variation of the irisin level in population. So we also evaluated non parametric method taking the median value of the corresponding control as cut-off. In this approach we observed there is significant lowering of irisin level in obese PCOS compared to corresponding obese healthy controls. However the non obese PCOS does not show any significant difference which could be explained by low demand of insulin in non obese patients leads to less chance of metabolic dysregulation.

Among the reproductive hormones only progesterone and testosterone showed significantly correlated with irisin level innonobese PCOS and obesePCOSpatients respectively. Similar observation was also reported previously however physiological interpretation of the relation yet to be evaluated^9^.

As like previous report in our study population also we did not observe fasting glucose level in diabetic range(70-100 mg/dL normal range of fasting glucose).however when we calculated HOMA-IR which combines the fasting glucose and insulin level, it showed relative insulin resistance (>3) in both obese and non obese PCOS as well as in obese healthy control. As irisin is functionallyrelated to increase insulin sensitivity^19,20^, the observation of relative insulin resistance in obese and non obese PCOS group is consistent with the observation of low irisin level in PCOS.

To evaluate theinterplay of insulin resistance with the irisin level in PCOS we determined the lower cut-off value of irisin in normal healthy individual and upper cut-off of insulin level in normal healthy non obese individual.Though there is significant increase of insulin level in low irisin (<14 ng/ml) group than in normal level irisin (>14 ng/ml) groupin both obese and non obese PCOS,however insulin resistance (>19.7 μU/ml) was statistically associated only with obese PCOS having low irisin level.As obese patients required high irisin demand to overcome the relative insulin resistance, the low irisin response in obese PCOS patients leads to insulin resistance. However in case of non obese subjects as the demand of irisin as well as insulin is low, even in lower level of irisin in non obese PCOS does not affect insulin sensitivity. The observation was also supported by previous report that low irisin is associated with insulin resistance and early Type II Diabetes Melitus^20,21^. Future prospective study is needed to evaluate frequency of conversion of type II Diabetes in the high risk (low irisin, obesity) PCOS group.

We conclude that a sub group of PCOS having low irisin level and obesity could be segregated as high risk PCOS who may have risk to develop T2DM. The high risk group could be followed-up more frequently for early detecting T2DM and or underwent any prophylactictherapywhich together may prevent many complication of the late stage disease. At the same time identifying this high risk group in preclinical stage will open up an avenue of therapeutic intervention by supplementing irisin.Further study is required with larger sample size to reproduce the observation with high statistical strength.

## Data availability

All the relevant Anonymized data will be available from the corresponding author upon reasonable request through data transfer agreements approved by the stakeholders,

## Acknowledgement

This work was supported by the Department of Health Research (DHR), Ministry of Health and Family Welfare (MOHFW), Government of India.

## Supplementary Materials

This PDF file includes:Table S1, Fig. S1

## Reference

1. Singh, S., Pal, N.; Shubham, S., Sarma, D.K., Verma, V., Marotta, F., Kumar, M. Polycystic Ovary Syndrome: Etiology, Current Management, and Future Therapeutics. J. Clin. Med. 2023, 12, 1454. 10.3390/jcm12041454

2. Bharali MD, Rajendran R, Goswami J, Singal K, Rajendran V. Prevalence of Polycystic Ovarian Syndrome in India: A Systematic Review and Meta-Analysis. Cureus. 2022 9;14(12):e32351.

3. Balen, A.H., Tan, S.L., MacDougall, J., Jacobs, H.S. Miscarriage rates following in-vitro fertilization are increased in women with polycystic ovaries and reduced by pituitary desensitization with buserelin. Hum. Reprod. 1993, 8, 959–964.

4. Azziz, R.; Woods, K.S.; Reyna, R.; Key, T.J.; Knochenhauer, E.S.; Yildiz, B.O. The prevalence and features of the polycystic ovary syndrome in an unselected population. J. Clin. Endocrinol. Metab. 2004, 89, 2745–2749.

5. Rotterdam ESHRE/ASRM-Sponsored PCOS consensus workshop group. Revised 2003 consensus on diagnostic criteria and long-term health risks related to polycystic ovary syndrome (PCOS).HumReprod. 2004;19:41–47.

6. Forslund M, Landin-Wilhelmsen K, Trimpou P, Schmidt J, Brännström M, Dahlgren E. Type 2 diabetes mellitus in women with polycystic ovary syndrome during a 24-year period: importance of obesity and abdominal fat distribution. Hum Reprod Open.;2020(1).

7. Bostrom P, Wu J, Jedrychowski MP, Korde A, Ye L, Lo JC, Rasbach KA, Boström EA, Choi JH, Long JZ, Kajimura S, Zingaretti MC, Vind BF, Tu H, Cinti S, Højlund K, Gygi SP, Spiegelman BM. A PGC1-α-dependent myokine that drives brown-fat-like development of white fat and thermogenesis. Nature. 2012;481:463–8.

8. Schumacher MA, Chnnam N, Ohashi T, Shah RS, Erickson HP. Structure of irisin reveals a novel inter subunit beta-sheet fibronectin (FNIII) dimer; implications for receptor activation. J Biol Chem 2013;288:33738–33744.

9. Sharan B, Sagili H, Kamalanathan SK, Lakshminarayanan S. Serum Irisin Levels and its Association with Blood Glucose and Insulin Indices in Diagnosing Insulin Resistance in Adolescents with Polycystic Ovarian Syndrome. J Hum Reprod Sci. 2021;14(2):137–143.

10. Abali R, Temel Yuksel I, Yuksel MA, Bulut B, Imamoglu M, Emirdar V, Unal F, Guzel S, Celik C. Implications of circulating irisin and Fabp4 levels in patients with polycystic ovary syndrome. J ObstetGynaecol. 2016 Oct;36(7):897–901. doi: 10.3109/01443615.2016.1174200. Epub 2016 May 16. PMID: 27184575.

11. Gao S, Cheng Y, Zhao L, Chen Y, Liu Y. The relationships of irisin with bone mineral density and body composition in PCOS patients. Diabetes Metab Res Rev. 2016;32(4):421–8.

12. Chia Lin Chang, Shang Yu Huang, Yung Kuei Soong, Po Jen Cheng, Chin-Jung Wang, I. Ting Liang, Circulating Irisin and Glucose-Dependent Insulinotropic Peptide Are Associated With the Development of Polycystic Ovary Syndrome, The Journal of Clinical Endocrinology & Metabolism, 2014;99 (12).

13. Li H, Xu X, Wang X, Liao X, Li L, Yang G, Gao L. Free androgen index and Irisin in polycystic ovary syndrome. J Endocrinol Invest. 2016 May;39(5):549–56.

14. Adamska A, Karczewska-Kupczewska M, Lebkowska A, Milewski R, Górska M, Otziomek E, Nikolajuk A, Wolczynski S, Kowalska I. Serum irisin and its regulation by hyperinsulinemia in women with polycystic ovary syndrome. Endocr J. 2016;63(12):1107–1112.

15. Li M, Yang M, Zhou X, Fang X, Hu W, Zhu W, Wang C, Liu D, Li S, Liu H, Yang G, Li L. Elevated circulating levels of irisin and the effect of metformin treatment in women with polycystic ovary syndrome. J Clin Endocrinol Metab. 2015;100(4):1485–93

16. Bostancı MS, Akdemir N, Cinemre B, Cevrioglu AS, Özden S, Ünal O. Serum irisin levels in patients with polycystic ovary syndrome. Eur Rev Med Pharmacol Sci. 2015;19(23):4462–8.

17. MedCalc Software Ltd. Comparison of proportions calculator. https://www.medcalc.org/calc/comparison_of_proportions.php (xVersion 22.014)

18. Luo, Y., Qiao, X., Xu, L. et al. Irisin: circulating levels in serum and its relation to gonadal axis. Endocrine, 2022; 75, 663–671.

19. Farhan FS, Hussien SS. Irisin as a Novel Marker for Insulin Resistance in Iraqi Women with Polycystic Ovary Syndrome Before and After Metformin Therapy. J ObstetGynaecol India. 2019 Oct;69(Suppl 2):194–200.

20. Shi, X., Lin, M., Liu, C. et al. Elevated circulating irisin is associated with lower risk of insulin resistance: association and path analyses of obese Chinese adults. BMC EndocrDisord 16, 44 (2016).

21. Akyuz A, Mert B, Ozkaramanli Gur D, Mucip Efe M, Aykac H, Alpsoy S, Guzel S. Association of lower serum irisin levels with diabetes mellitus: Irrespective of coronary collateral circulation, and syntax score. North Clin Istanb. 2021;8(6):607–614.

