## Supplemental Table S1, Supplemental Figure S1 for "Interplay of Low Serum Irisin Level and Insulin Resistance in Obese Polycystic Ovary Syndrome Patients: Potential Biomarker for Pre-clinical Risk Assessment and therapeutic intervention"

**TableS1: Correlation of serum irisin with anthropometric and reproductive hormonal profile of PCOS patients including obese and non obese**

| **S.No.** | **Parameter** | **Obese PCOS Patients** | | **Non obese PCOS Patients** | |
| --- | --- | --- | --- | --- | --- |
|  |  | **Correlation coefficient (R^2)** | ***p*** | **Correlation coefficient (R^2)** | ***p*** |
| a. | Testosterone | 0.4194 | 0.002* | 0.0883 | 0.2030 |
| b. | Progesterone | 0.0167 | 0.5871 | 0.9436 | <0.0001* |
| c. | Prolactin | 0.1869 | 0.057 | 0.0113 | 0.060 |
| d. | Insulin | 0.0596 | 0.2617 | 0.1146 | 0.0908 |
| e. | Fasting blood sugar | 0.001 | 0.8503 | 0.0010 | 0.8774 |
| f. | Age | 0.0596 | 0.2195 | 0.004 | 0.7568 |
| g. | BMI | 5.743 | 0.9905 | 0.0002 | 0.9415 |
| h. | W/H ratio | 0.0325 | 0.3678 | 0.0068 | 0.6815 |
| i. | AC | 0.0033 | 0.7736 | 0.0004 | 0.9128 |
| j. | Menarche | 0.0027 | 0.7936 | 0.0215 | 0.4651 |
| k. | LH | 0.0043 | 0.7450 | 0.0195 | 0.4873 |
| l. | FSH | 0.0318 | 0.3732 | 0.0208 | 0.4727 |
| m. | TSH | 0.0068 | 0.6818 | 0.0487 | 0.2686 |

BMI=Body Mass Index, W/H ratio=Waist-Hip Ratio (WHR),AC**=**Abdominal Circumference, LH=Luteinizing hormone, FSH=follicle-stimulating hormone, TSH=Thyroid-stimulating hormone

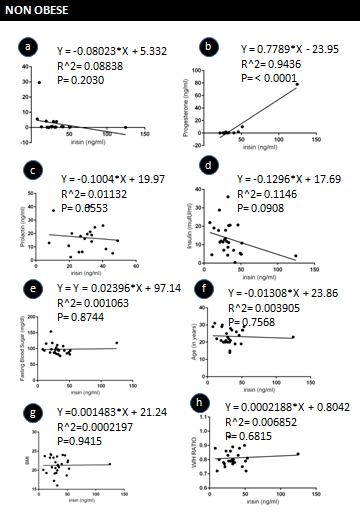

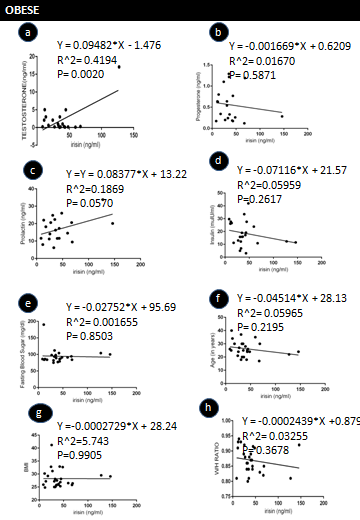

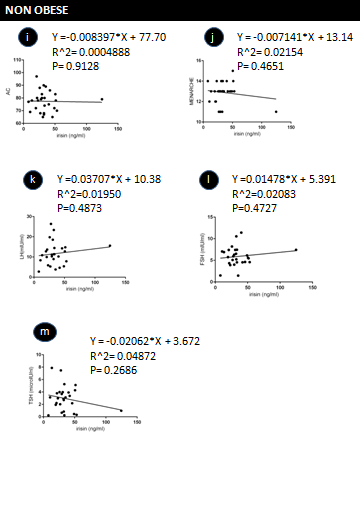

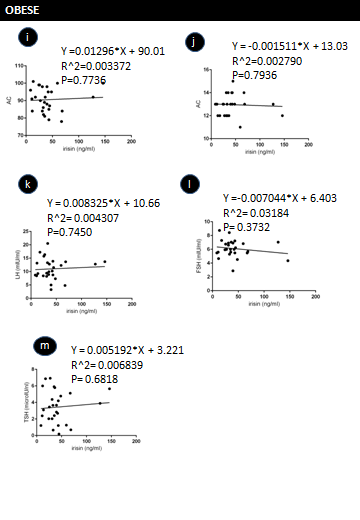

Figure S1: Correlation between Serum irisin level and Reproductive hormonal profile as well as demographic profile in non obese PCOS patients: a. testosterone; b. Progesterone .Prolactin; insulin; e. blood sugar; f. age; g. BMI; h. W/H ratio ; i. AC; j. menarche; k. LH; l. FSH; m. TSH
